# Analysis of Single and Polydrug Profiles in the All of Us Cohort: Self-Reported Health, Wearable Sleep Metrics, and Family History

**DOI:** 10.1101/2025.10.31.25339286

**Authors:** Akane Sano, Nidal Moukaddam

## Abstract

Substance use remains a major public health concern in the United States, with diverse patterns of single- and multi-drug use linked to varying health outcomes across populations. Leveraging multimodal data from the All of Us Research Program, including survey responses, electronic health records, and Fitbit-derived metrics, this study examines associations between drug use categories and indicators of general health, sleep quality, physical activity, and family history of substance use. Polydrug and stimulant users reported significantly lower health ratings, greater sleep fragmentation, and reduced activity levels, while cannabis-only users showed comparable outcomes to non-users. Elevated family history among polydrug users supports hypotheses of intergenerational risk transmission. Wearable-derived sleep metrics emerged as sensitive indicators of substance-related health disparities and underscore the value of passive monitoring in addiction research. These findings highlight the potential of integrated behavioral data to inform personalized prevention strategies and scalable digital health interventions.

## 1 Introduction

Substance use and its health consequences remain a critical public health issue in the United States, contributing to increased morbidity, mortality, and healthcare burden [14]. Patterns of use, including single-drug use, polysubstance use, and abstention, vary widely across sociodemographic groups and life stages, shaped by factors such as access, stigma, trauma, and familial exposure [7]. While national surveys provide broad prevalence estimates, they often lack the granularity needed to explore how substance use behaviors intersect with day-to-day health experiences, especially in relation to sleep, physical activity, and psychosocial functioning [13].

This study leverages the All of Us Research Program’s uniquely multimodal dataset [15], which includes person-level survey responses, electronic health records (EHR), and biometric data from wearable devices. This integration allows us to examine how drug use patterns correlate with behavioral and health indicators across multiple domains. Specifically, we assess whether participants who report single- or multi-drug use differ significantly from non-users in general health ratings, sleep measures, and family history of substance use.

We hypothesize that individuals reporting multi-drug use will show lower general health ratings and poorer sleep quality compared to single-drug users and non-users. We also expect family history of substance use to be more prevalent among multi-drug users, suggesting potential intergenerational transmission of risk [12, 11]. These hypotheses are grounded in prior research linking polysubstance use to disrupted circadian rhythms, elevated stress, and diminished self-rated health [9, 4, 17].

By incorporating wearable-derived metrics, such as sleep duration and efficiency, this study addresses a key gap in addiction science: the lack of objective, continuous behavioral data to complement self-report and clinical records. Through this multimodal approach, we aim to uncover nuanced patterns in substance use and everyday health, with implications for early intervention, digital health monitoring, and scalable, personalized prevention strategies [5, 2, 6].

## 2 Methods

### 2.1 Cohort Selection and Data Sources

This study utilized data from the All of Us Research Program, a nationwide initiative designed to advance precision medicine through diverse, longitudinal health data. Participants were included if they completed the substance use survey (**concept ID: 1333017: past month drug use**). All data, including survey responses, electronic health records (EHR), and wearable metrics, were accessed via the All of Us Researcher Workbench and analyzed in a secure JupyterLab environment.

### 2.2 Demographic Information

Demographic variables were derived from core surveys and EHR records. Age was calculated at the time of survey completion. Race and ethnicity were harmonized using standardized categories from the All of Us Research Program. Race was self-reported and included: White, Black or African American, Asian, American Indian or Alaska Native, Middle Eastern or North African, Native Hawaiian or Other Pacific Islander, None of these, I prefer not to answer, and PMI: Skip. Ethnicity was categorized separately as Hispanic or Latino, Not Hispanic or Latino, I prefer not to answer, or PMI: Skip.

### 2.3 Drug Use Categorization

Participants were classified into mutually exclusive drug use groups, (1) Single Drug Use, (2) Multiple Drug Use, and (3) No Use, based on self-reported survey responses about past month drug use.

Participants were classified into mutually exclusive drug use groups based on survey responses. Most categories such as Cannabis Only, Opioids Only, and Cocaine Only, reflect single-substance use. Polydrug Use includes individuals reporting three or more substances, representing a high-risk group [4]. Other Combination captures less common pairings not covered by predefined dyads (e.g., cannabis + opioids).

Group definitions prioritized interpretability, clinical relevance, and statistical power. No Use served as the baseline. Cannabis Only was isolated due to its prevalence and distinct risk profile [1]. Prescription-based groups (e.g., Opioids Only, Sedatives Only, Stimulants Only) reflected medically relevant patterns [10], while high-risk substances like Methamphetamine Only and Cocaine Only were retained as standalone categories [8]. Intermediate groups (e.g., Cannabis + One Other, Opioids + Sedatives) enabled nuanced co-use analysis [16].

### 2.4 Drug Use Related Measures

Additional variables included family history of drug use and age at diagnosis. Family history was quantified by counting mentions across relevant survey responses (concept ID: 836851), providing a proxy for intergenerational risk. Age at diagnosis (concept ID: 1384514) was derived from the earliest recorded drug use disorder diagnosis and used as an indicator of early health burden.

### 2.5 Health Indicators

Five self-reported health indicators were extracted using OMOP concept IDs: General Health Rating (1585711), Quality of Life Rating (1585717), Physical Health Interference (1585723), Mental Health Interference (1585729), and Social Satisfaction (1585735). Each response was mapped to standardized categories such as “Excellent,” “Fair,” or “Very Dissatisfied” to facilitate interpretation and analysis.

### 2.6 Fitbit-Derived Metrics

Daily sleep data were aggregated per participant. Metrics included minutes asleep, minutes in bed, minutes awake, minutes restless, and minutes spent in deep, light, or REM sleep. Additional indicators such as Sleep efficiency, fragmentation, and REM ratio were computed. Sleep efficiency is calculated as minutes asleep divided by minutes in bed; fragmentation combines minutes awake and restless; REM ratio is the proportion of REM sleep relative to total sleep time.

### 2.7 Analytical Approach

Drug use groups were compared across health indicators and Fitbit-derived metrics using a combination of statistical approaches. For continuous variables such as age and sleep efficiency, Kruskal-Wallis tests were employed to assess overall group differences. When significant results were observed, Tukey HSD post hoc tests were conducted to identify specific pairwise differences between drug use categories. Categorical health indicators, including self-reported general health and quality of life ratings, were analyzed using chi-square tests. To further interpret these associations, standardized residuals were calculated to highlight group-category pairs that contributed most strongly to the observed significance.

## 3 Results

### 3.1 Drug Use Distribution

Participants were classified into mutually exclusive drug use groups (Table 1, No Use (68%), Single Drug Use (20%), and Multiple Drug Use (13%) based on survey responses. The No Use group was the most prevalent and served as the reference category.

**Table 1:** Participant Distribution by Drug Use Category (Past Month Drug Use)

| Category | Count | % |
| --- | --- | --- |
| No Used | 34,813 | 68.0 |
| Single Drug | 10,391 | 20.3 |
| Multiple Drugs | 6,600 | 12.9 |

Among single-drug users, the most commonly reported substances were cannabis (0.89%), prescription sedatives (0.50%), and PMI: Skip (1.03%), indicating missing or ambiguous responses. Use of prescription opioids (0.20%) was less common, and other substances such as stimulants, hallucinogens, and methamphetamine each accounted for less than 0.01%.

In contrast, multiple-drug users showed broader and more intensive substance use. Higher proportions reported cannabis (1.53%), sedatives (1.31%), and opioids (0.61%), along with elevated use of prescription stimulants (0.44%), hallucinogens (0.07%), and cocaine (0.05%) (Table 2). The PMI: Skip category remained notable (0.45%), suggesting some degree of non-disclosure or incomplete reporting. These patterns highlight the predominance of cannabis and sedative use across both groups, while underscoring the greater diversity and intensity of substance use among multiple-drug users.

**Table 2:**
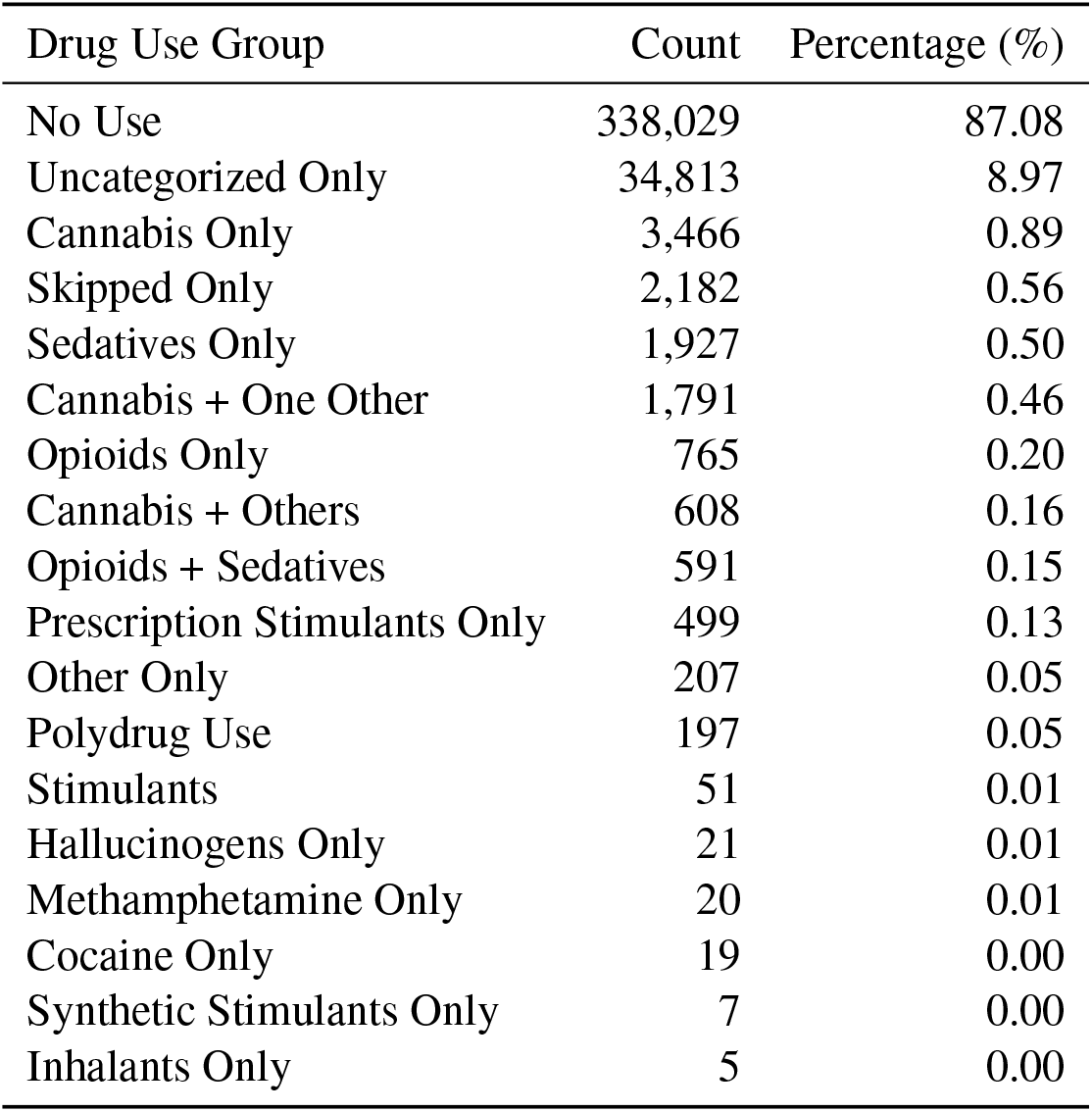
Drug Use Group Counts and Percentages.

Among participants reporting a drug use disorder, the majority were first diagnosed in adulthood (18–64 years): 71.4% of polydrug users, 81.6% of single-drug users, and 75.8% of those in the “No Used” group. Adolescent-onset diagnoses (12–17 years) were more common among polydrug users (25.0%) than single-drug users (18.4%), with 23.1% of “No Used” participants also reporting adolescent diagnoses likely reflecting historical use. Childhood (0–11 years) and older adult (65–74 years) diagnoses were rare across all groups (<2.5%). Missing responses (PMI: Skip) were minimal. These patterns suggest earlier onset among polydrug users, underscoring the need for targeted screening and early intervention.

Table 3 shows how often participants in each drug use category reported a family member with a drug use disorder. Most responses were PMI: Skip, especially among single drug users (82.5%), suggesting low disclosure. Among those who answered, siblings and self were the most common mentions, particularly in the multiple drug group. Parental mentions (mother, father) were also more frequent in that group, hinting at possible familial patterns.

**Table 3:** Family Role Mentions by Drug Use Group (%)

| Role | No | Single | Multiple |
| --- | --- | --- | --- |
| PMI: Skip | 63.9 | 82.5 | 71.2 |
| Sibling | 11.0 | 6.2 | 8.9 |
| Self | 8.1 | 2.5 | 6.7 |
| Father | 5.3 | 2.1 | 3.9 |
| Mother | 4.6 | 2.1 | 3.4 |
| Son | 3.2 | 2.3 | 2.8 |
| Daughter | 2.0 | 1.3 | 1.6 |
| Grandparent | 2.0 | 0.9 | 1.6 |

### 3.2 Race, Ethnicity, and Age

Race composition was broadly similar across drug use groups, with most participants identifying as White, Black, or Asian. Multiple drug use was slightly more common among Middle Eastern/North African (2.8%) and Asian (2.0%) individuals. Age differed by group: single drug users were oldest (mean = 64.4 years), followed by multiple drug users (60.5), and non-users (57.6).(Kruskal-Wallis H = 358,411.79, p < 0.0001). Ethnicity did not differ significantly across groups (chi-square = 0.00, p = 1.00).

### 3.3 Health and Wellbeing Indicators

Participants who reported Single Drug Use consistently showed the lowest ratings across general health (Table 4), quality of life, and social satisfaction. For example, 37.9% rated their general health as “Fair” and only 2.9% selected “Excellent”. Similarly, 25.1% rated their quality of life as “Good”, while just 3.4% reported “Excellent” social satisfaction. These individuals also reported elevated physical and mental health interference, with 10.8% indicating that mental health interfered “Very much” with daily life.

**Table 4:** General Health Rating by Drug Use Group (%)

| Rating | No | Single | Multiple |
| --- | --- | --- | --- |
| Skipped | 1.0 | 0.6 | 0.5 |
| Poor | 10.9 | 12.4 | 8.5 |
| Fair | 30.9 | 37.9 | 31.0 |
| Good | 34.0 | 32.2 | 33.6 |
| Very Good | 18.7 | 14.2 | 20.7 |
| Excellent | 4.6 | 2.9 | 5.6 |

Multiple Drug Users reported broader health challenges, with 23.5% experiencing “Very much” mental health interference and 6.6% reporting “Very much” physical health interference. Despite this, they showed slightly higher ratings in the upper categories—20.7% selected “Very Good” and 5.6% “Excellent” for general health, and 17.0% and 7.7% did so for social satisfaction, suggesting a more resilient profile.

In contrast, No Used participants generally reported the most favorable outcomes. A combined 57.3% rated their general health as “Good”, “Very Good”, or “Excellent”, and only 14.0% reported “Very much” mental health interference. These patterns underscore the gradient of self-rated well-being across drug use groups and highlight the multidimensional burden associated with substance use.

Chi-square tests revealed significant associations between drug use category and both general health ratings and social satisfaction. Polydrug and stimulant users were more likely to report poor health and lower social satisfaction, while cannabis-only users tended to report better health outcomes. Other health indicators such as quality of life, physical and mental health interference could not be reliably tested due to limited variation or sample size.

### 3.4 Sleep Patterns

Kruskal-Wallis tests revealed significant differences in wearable-derived health metrics across drug use groups. Sleep efficiency was significantly lower among participants in the Polydrug Use group (H = 18.3, p < 0.001), while sleep fragmentation was elevated among those reporting stimulant and opioid use (H = 21.7, p < 0.001). Post-hoc Tukey HSD comparisons identified significant pairwise differences between Cannabis Only and Polydrug Use groups, as well as between No Use and Stimulant users, highlighting distinct behavioral health profiles associated with specific substance use patterns.

## 4 Discussion and Conclusions

This analysis of All of Us survey, EHR, and Fitbit data revealed consistent associations between substance use patterns and multiple dimensions of health and behavior. Participants classified as polydrug or stimulant users reported notably lower general health ratings, greater sleep fragmentation, and reduced physical activity compared to single-drug users and non-users. For example, 23.5% of polydrug users reported that mental health interfered “very much” with daily life, and their sleep efficiency was significantly lower than other groups. These findings align with prior research linking polysubstance use to heightened physiological and psychological burden [4, 3], and suggest that behavioral metrics derived from wearables may serve as sensitive indicators of underlying health disparities in substance-using populations [2, 6].

Elevated family history of drug use among polydrug and stimulant users further supports the hypothesis of intergenerational risk transmission. While causal inference is limited by the cross-sectional design, the strength and consistency of these associations underscore the importance of incorporating familial context into addiction risk models [12, 11].

Notably, cannabis-only users did not differ significantly from non-users across most health and behavioral metrics, suggesting that isolated cannabis use may carry a lower health burden than more complex substance use profiles. However, intermediate patterns observed in *Cannabis + One Other* and *Cannabis + Others* groups indicate that even modest co-use may influence health trajectories, with slight increases in sleep fragmentation and reduced physical activity.

The integration of Fitbit-derived metrics added a valuable behavioral layer to the analysis. Sleep efficiency and sleep fragmentation emerged as particularly sensitive to drug use category, with polydrug users exhibiting the most disrupted sleep profiles. These findings echo emerging literature on the role of sleep dysregulation in addiction vulnerability and recovery [9, 13], and highlight the potential of passive monitoring tools to identify at-risk individuals or track treatment progress.

Several limitations should be acknowledged. First, the All of Us dataset is observational and relies on self-reported measures, which may introduce bias in drug use disclosure and health ratings. Second, Fitbit data were available for a subset of participants, and wear-time compliance may vary. Third, the cross-sectional nature of the analysis precludes temporal or causal interpretations. Additionally, rare drug categories such as methamphetamine-only or cocaine-only had limited sample sizes, reducing statistical power and generalizability.

The clinical significance of these findings needs to be further explored. Cannabis has long been viewed as a gateway drug, with an implicit perception that it is milder than other drugs. This is not necessarily accurate with the increase in THC potency and with the advent of synthetic cannabinoids. It is also possible that polysubstance users have predisposing factors such as worse impulsivity, mood disorders and concomitant trauma that shapes their substance use. Specifically, it is unknown whether a marked preference for a specific substance (being more selective) may reflect less severe addiction in general, less “desperation” to numb negative feelings, etc. those are intriguing avenues to pursue in our quest to understand substance use correlates.

Despite these constraints, the findings demonstrate the feasibility and value of integrating survey, clinical, and wearable data to characterize substance use patterns and their health correlates. Future research could build on this framework by incorporating longitudinal data to examine transitions between drug use categories, linking behavioral metrics to treatment engagement or relapse, and developing predictive models using multimodal inputs. Such approaches may inform personalized prevention strategies and adaptive interventions [2].

In conclusion, this study provides a multidimensional snapshot of how substance use behaviors intersect with self-rated health, family history, and real-world behavioral patterns. The results reinforce the importance of distinguishing between single- and multi-drug use in addiction science and suggest that wearable data may offer a scalable, non-invasive tool for monitoring health in substance-using populations. As digital health technologies become more integrated into clinical care, these insights may help guide more responsive, individualized approaches to addiction prevention and treatment.

## Data Availability

NIH All of US program

